# Arrhythmia and Survival Outcomes among Black and White Patients with a Primary Prevention Defibrillator

**DOI:** 10.1101/2023.05.01.23289362

**Authors:** Arwa Younis, Sanah Ali, Eileen Hsich, Ido Goldenberg, Scott McNitt, Bronislava Polonsky, Mehmet K. Aktas, Valentina Kutyifa, Oussama M. Wazni, Wojciech Zareba, Ilan Goldenberg

## Abstract

**Background:** Black Americans have a higher risk of non-ischemic cardiomyopathy (NICM) than White Americans. We aimed to evaluate racial disparities in the risk of tachyarrhythmias among patients with an implantable cardioverter defibrillator (ICD).

**Methods:** The study population comprised 3,895 ICD recipients enrolled in the U.S. in primary prevention ICD trials. Outcome measures included first and recurrent ventricular tachy-arrhythmia (VTA) and atrial tachyarrhythmia (ATA), derived from adjudicated device data, and death. Outcomes were compared between self-reported Black vs. White patients with a cardiomyopathy (ischemic [ICM] and NICM).

**Results:** Black patients were more likely to be female (35% vs 22%) and younger (57±12 vs 62±12) with a higher frequency of comorbidities. Blacks patients with NICM compared with Whites patients had a higher rate of first VTA, fast VTA, ATA, appropriate-, and inappropriate-ICD-therapy (VTA≥170bpm: 32% vs. 20%; VTA≥200bpm: 22% vs. 14%; ATA: 25% vs. 12%; appropriate 30% vs 20%; and inappropriate: 25% vs. 11%; p<0.001 for all). Multivariable analysis showed that Black patients with NICM experienced a higher risk of all types of arrhythmia/ICD-therapy (VTA≥170bpm: HR=1.69; VTA≥200bpm: HR=1.58; ATA: HR=1.87; appropriate: HR=1.62; and inappropriate: HR=1.86; p≤0.01 for all), higher <u>burden</u> of VTA, ATA, ICD therapies, and a higher risk of death (HR=1.86; p=0.014). In contrast, in ICM, the risk of all types of tachyarrhythmia, ICD therapy, or death was similar between Black and White patients.

**Conclusions:** Among NICM patients with an ICD for primary prevention, Black compared with White patients had a high risk and burden of VTA, ATA, and ICD therapies.

**Clinical Perspective:** *What Is New?:*

- Black patients have a higher risk of developing non-ischemic cardiomyopathy (NICM) but are under-represented in clinical trials of implantable cardioverter defibrillators (ICD). Therefore, data on disparities in the presentation and outcomes in this population are limited.
- This analysis represents the largest group of self-identified Black patients implanted in the U.S. with an ICD for primary prevention with adjudication of all arrhythmic events.

*What Are the Clinical Implications?:*

- In patients with a NICM, self-identified Black compared to White patients experienced an increased incidence and burden of ventricular tachyarrhythmia, atrial tachyarrhythmia, and ICD therapies. These differenced were not observed in Black vs White patients with ischemic cardiomyopathy (ICM).
- Although Black patients with NICM were implanted at a significantly younger age (57±12 vs 62±12 years), they experienced a 2-fold higher rate of all-cause mortality during a mean follow up of 3 years compared with White patients.
- These findings highlight the need for early intervention with an ICD, careful monitoring, and intensification of heart failure and antiarrhythmic therapies among Black patients with NICM.

## Introduction

In the United States (U.S.), heart failure (HF) disproportionately affects Black patients, also known as African-Americans, a population in whom the prevalence of the disease is higher, more often caused by non-ischemic cardiomyopathy (NICM), with a more severe disease trajectory when compared with White patients.^1–3^ Those disparities are less evident in patients with ischemic cardiomyopathy (ICM).^4–6^

There are conflicting findings when looking at outcomes of Black patients with HF. Some studies have suggested no significant differences in patient-reported health status between Black and White HF patients,^7, 8^ whereas, others suggest that Black patients experience significantly worse clinical outcomes, including cardiovascular mortality and readmissions.^9, 10^ These conflicting results are also evident when assessing the risk of ventricular tachyarrhythmia (VTA) and/or implantable cardioverter defibrillator (ICD) therapies among Black patients. Several reports have shown that Black patients with HF experience increased risk of developing VTA, as well as sudden cardiac death (SCD), compared to White patients.^8, 9^ On the other hand, several studies reported a similar VTA risk between Black and White patients.^7, 8^

Black patients are underrepresented in all major ICD randomized clinical trials, generally accounting for less than 20% of a study cohort which may be contributing to inconsistent racial differences in outcomes. Additionally, implantation of an ICD has been shown to be significantly less common among Blacks compared to White patients, especially for primary prevention, despite data suggesting increased risk of SCD.^8, 9^ Importantly, none of the aforementioned studies focused on NICM, which is more prevalent and with a more severe HF trajectory among Black compared to White patients possibly due to associated comorbidities, such as diabetes and hypertension.^11–15^ Thus, ICD candidacy for primary prevention does not consider potential differences in the risk of arrhythmic events, appropriate and inappropriate device therapy, and survival among Black and White patients based on type of cardiomyopathy.

In this study we aimed to assess differences in arrhythmic and survival outcomes between Black and White patients according to cardiomyopathy type among patients enrolled in the U.S. landmark ICD trials, with a focus on the risk of; 1) first and recurrent VTA; 2) Device detected atrial tachyarrhythmias (ATA); 3) appropriate and inappropriate ICD therapy; and 4) all-cause mortality. Outcomes were assessed separately in patients with NICM and ICM.

## Methods

### Study Population

The study cohort consisted of 3,895 United States (U.S,) patients with an ICD or cardiac resynchronization therapy-defibrillator (CRT-D) who were enrolled in five primary prevention ICD studies conducted between July 1997 and January 2017. Design, study protocol, and results of each of these trials have been published previously. Briefly, MADIT-II and MADIT-Risk enrolled a total of 1130 patients implanted with an ICD for primary prevention therapy due to ICM. Inclusion criteria were mainly prior myocardial infarction and a left ventricular ejection fraction (LVEF) of ≤30%.^16^ MADIT-CRT enrolled 1,814 HF patients (N=993 ICM and N=821 NICM patients), an LVEF of 30% or less, a QRS≥130 milliseconds, and New York Heart Association (NYHA) functional class I/II. Patients were randomly assigned in a 3:2 ratio to receive CRT-D (N=1,083) or an ICD alone (N=731).^17^ MADIT-RIT enrolled 1,465 patients (N=838 ICM and N=627 NICM patients) and all patients met guideline criteria to receive an ICD or CRT-D for primary prevention.^18^ The RAID trial enrolled 1,012 high-risk ICD patients (primary prevention N=669, secondary prevention N=343) who were randomized to treatment with ranolazine or placebo. Treatment with ranolazine did not affect the VTA risk or the all-cause mortality risk significantly. For the current study we included primary prevention RAID patients only (N=669).^19^

For the primary analysis, the total study cohort was stratified by NICM and ICM. Outcomes in each type of heart disease cohort were compared between self-identified Black and White patients. The study design and population are summarized in the **Supplemental Figure 1**.

Each study was approved by the institutional review boards at the respective enrolling sites before participation. All patients provided informed consent before enrollment into the respective trials.

### Definitions and End Points

In all MADIT trials and in RAID, device interrogations were reviewed and adjudicated blindly by an independent committee of at least two experienced electrophysiologists and cardiologists using identical definitions. Details of device programming and VTA definitions are provided in **Table 1** of the **Supplementary Appendix**. In all studies ICD programming were similar for Black and White.

**Table 1.** – Baseline Characteristics of the Study Cohort by Cardiomyopathy BMI= body mass index; BP= blood pressure; bpm= beat per minute; CRT-D= cardiac resynchronization therapy with a defibrillator; LBBB= left bundle branch block; NYHA= New York Heart Association class; CABG= coronary artery bypass grafting; ACE=angiotensin-converting enzyme; ARB=angiotensin II receptor blocker.

|  | **Ischemic** |  |  | **Non-ischemic** |  |  |
| --- | --- | --- | --- | --- | --- | --- |
| **Clinical Characteristics** | **White**<br>**(N=2276)** | **Black**<br>**(N=290)** | **p-value** | **White**<br>**(N=981)** | **Black**<br>**(N=348)** | **p-value** |
| Age (years) | 66±10 | 62±11 | <0.001 | 62±12 | 57±12 | <0.001 |
| Female | 326(14) | 90(31) | <0.001 | 380(39) | 155(45) | 0.058 |
| Hispanic Ethnicity | 66(3) | 6(2) | 0.45 | 34(3) | 1(0) | 0.002 |
| BMI, kg/m<sup>2</sup> | 29±6 | 29±6 | 0.42 | 30±7 | 32±8 | <0.001 |
| Systolic BP, mmHg | 123±18 | 125±20 | 0.021 | 122±17 | 123±19 | 0.031 |
| Diastolic BP, mmHg | 71±11 | 75±11 | <0.001 | 72±11 | 75±12 | <0.001 |
| Heart Rate, bpm | 70±12 | 72±12 | <0.001 | 71±12 | 75±13 | <0.001 |
| Ejection fraction, % | 26±6 | 25±7 | 0.022 | 25±7 | 23±8 | 0.021 |
| Ejection fraction ≤25% | 1189(52) | 165(57) | 0.14 | 576(59) | 244(70) | <0.001 |
| CRT-D | 643(28) | 60(21) | <0.001 | 592(60) | 156(45) | <0.001 |
| LBBB | 536(32) | 44(25) | 0.047 | 540(69) | 97(45) | <0.001 |
| QRS | 138±31 | 131±33 | 0.019 | 158±22 | 149±24 | <0.001 |
| NYHA ≥III | 1699(75) | 247(87) | <0.001 | 929(96) | 330(96) | 0.78 |
| Hypertension | 1526(67) | 247(85) | <0.001 | 585(60) | 283(82) | <0.001 |
| Atrial Arrhythmia* | 450(20) | 51(18) | 0.34 | 189(20) | 63(19) | 0.63 |
| Diabetes | 806(36) | 131(45) | <0.001 | 211(22) | 132(38) | <0.001 |
| Current Cigarette Use | 342(15) | 47(17) | 0.45 | 112(12) | 43(14) | 0.37 |
| Prior CABG | 1271(56) | 105(36) | <0.001 | 6(1) | 3(1) | 0.76 |

#### First arrhythmic event endpoints

The primary endpoint of the current study was the first occurrence of VTA, defined as ICD-recorded, treated or monitored, sustained ventricular tachycardia (VT) ≥170 bpm or ventricular fibrillation (VF). Secondary endpoints included: a) fast VTA ≥ 200 bpm b) any ATA event (atrial fibrillation or flutter [AF], or supraventricular tachycardia [SVT]), c) appropriate ICD therapy, and d) inappropriate ICD therapy.

#### Arrhythmic burden endpoints

Additional analysis was performed to assess the burden of all types of arrhythmia listed above (in a recurrent event analysis). Arrhythmic burden was defined as the total number of events per patient during follow-up. Analysis was limited to 10 events per patient to lessen the impact of subjects with large counts of arrhythmic occurrences.

Arrhythmias were defined by the adjudication committee based on combined evaluation of frequency, QRS morphology, and regularity of the rhythm. A VTA event was defined as lasting > 30 seconds, or if therapy was administered and terminated the arrhythmia (including polymorphic VT). Detection zone was ≥ 145 bpm in all studies. Arrhythmia events detected by the ICD and CRT-D devices were categorized as ATA if they included the occurrence of AF, or other supraventricular tachyarrhythmias. Regular SVT included regular atrial tachycardias, such as atrioventricular reentry tachycardia, and atrial tachycardia with 1:1 conduction. All devices in the current study were at least dual-chamber devices (1 electrode in the right atrium and 1 in the right ventricle). Inappropriate therapies due to non-arrhythmic causes (lead malfunction, device under sensing etc.) were excluded.

#### Mortality outcomes

To account for the competing risk of death we evaluated the following mortality outcomes: all-cause mortality; and non-arrhythmic mortality (defined as death without experiencing sustained fast VTA at any time during follow-up after ICD implantation).

### Statistical Analysis

Categorical data are summarized as frequencies and percentage in brackets (%). Continuous data are summarized as means ± standard deviations. Baseline characteristics were compared between the Black and White subgroups using Wilcoxon rank-sum test (for continuous variables) and Chi-square test (for categorical variables) in each cardiomyopathy separately.

Kaplan-Meier cumulative probabilities were used to graphically display the relationship of Black with the endpoints over time. The log-rank test was used for determination of statistical significance.

Multivariable Cox proportional hazards regression analysis was used to identify and evaluate the impact of Black on the risk of VTA and the other relevant endpoints. For the multivariable adjustment, relevant clinical covariates were determined based on the results of a stepwise selection process and were included if statistically significant at p<0.05 in multivariable models for either endpoint (VTA or death). Potential risk factor candidates are shown in the **Supplementary Table 2**. We used dichotomized functional form of continuous variables and continuous variable. Thresholds for categorization of numeric variables were pre-specified using clinically well-accepted criteria.

The following independent predictors were identified: age, sex, smoking, diastolic blood pressure, CRT-D, NYHA class, history of HF hospitalization, and history of ATA. These predictors were included in the multivariate model. All models were further adjusted for study type (MADIT II, RISK, CRT, RIT, and RAID) and study arm (treatment arm) by stratification.

Anderson Gill non-gap-time model was used to estimate the hazard ratios for *recurrent* VTA events, *recurrent* fast VTA ≥200 bpm, *recurrent* appropriate ICD shocks, and *recurrent* appropriate ICD ATPs. Models were adjusted for covariates as per above. Mean cumulative event rate was presented using the Ghosh-Lin curves to display mean number of recurrent events per patient as appropriate.

Within each group (Black and White) we used cumulative incidence function (CIF) curves to estimate the probability of first VTA, as the event of interest, and the probability of death without prior VTA as a competing risk.

Analysis in a total population model showed a statistically significant interaction in the association of Black compared with White patients with the risk of the primary endpoint by the type of cardiomyopathy ([ICM/NICM] p-value for interaction = 0.0083). Thus outcomes herein are reported separately for patients with NICM and ICM.

All tests of significance were two-tailed with a p-value<0.05 accepted to indicate statistical significance. Analysis was carried out using SAS statistical (version 9.4, SAS Institute, North Carolina).

## Results

### Baseline Characteristics

Among all ICD recipients, there were 638 (16%) self-identified Black patients and 3,257 White patients (84%). Mean age was 59±11 years and 245 (38%) were female. Black patients were more likely to have a NICM compared with ICM (26% vs. 11%; p<0.001) whereas White patients were more likely to have an ICM compared with NICM (p<0.001). Baseline characteristics, comparing Black and White patients with NICM and ICM are shown in **Table 1**. In both types of heart disease, self-identified Black compared to White patients were significantly younger (ICM: 62±11 vs. 66±10; NICM: 57±12 vs. 62±12; p<0.001 for each comparison), more likely to be female, and exhibited a higher frequency of cardiovascular comorbidities, including hypertension and diabetes (p<0.001 for all comparisons to White patients). Importantly, history of prior ATA or guidelines directed medical therapy (GDMT) was similar between Black and White patients except Black patients were more likely to be treated with aldosterone antagonists and other diuretics in both groups of ICM and NICM patients (**Table 1**).

### Patients with non-ischemic cardiomyopathy

#### First event endpoints

At 3 years, the Kaplan Meier cumulative probability of a first VTA (defined as VT≥170 bpm or VF) was significantly higher among Black than White patients (34% vs. 20%, respectively; p<0.001 for the overall difference during follow-up; Figure 1A). Fast VTA (defined as VT≥200 bpm or VF) occurred at a significantly higher rate among Blacks when compared with White patients (fast VTA: 24% vs. 15%; Figure 1B), as well as appropriate ICD therapy (30% vs 20%, respectively; p<0.001) and inappropriate ICD therapy (25% vs 11%, respectively; p<0.001) (**Supplementary Figures 2A-B, respectively**).

**Figure 1A-B.**
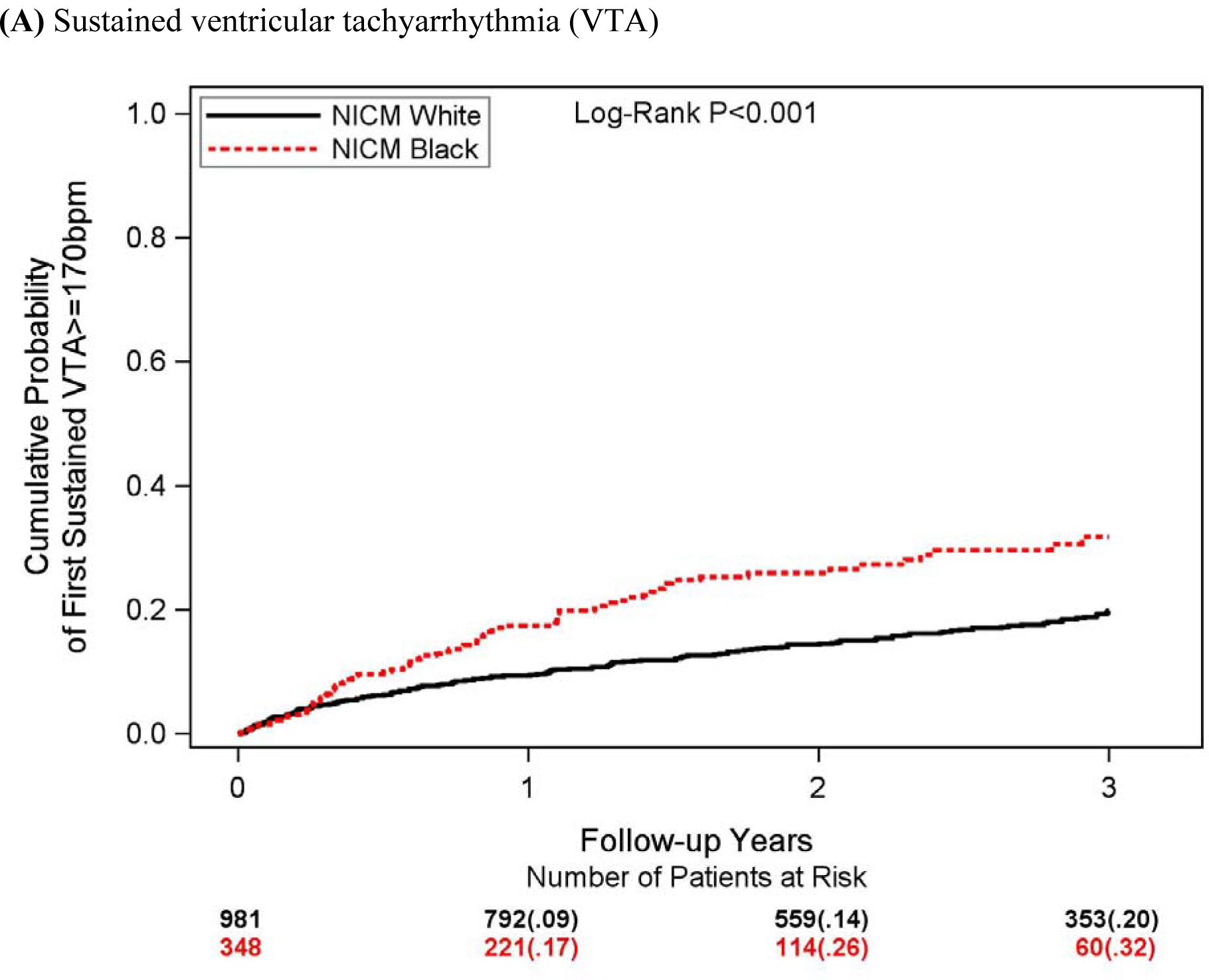
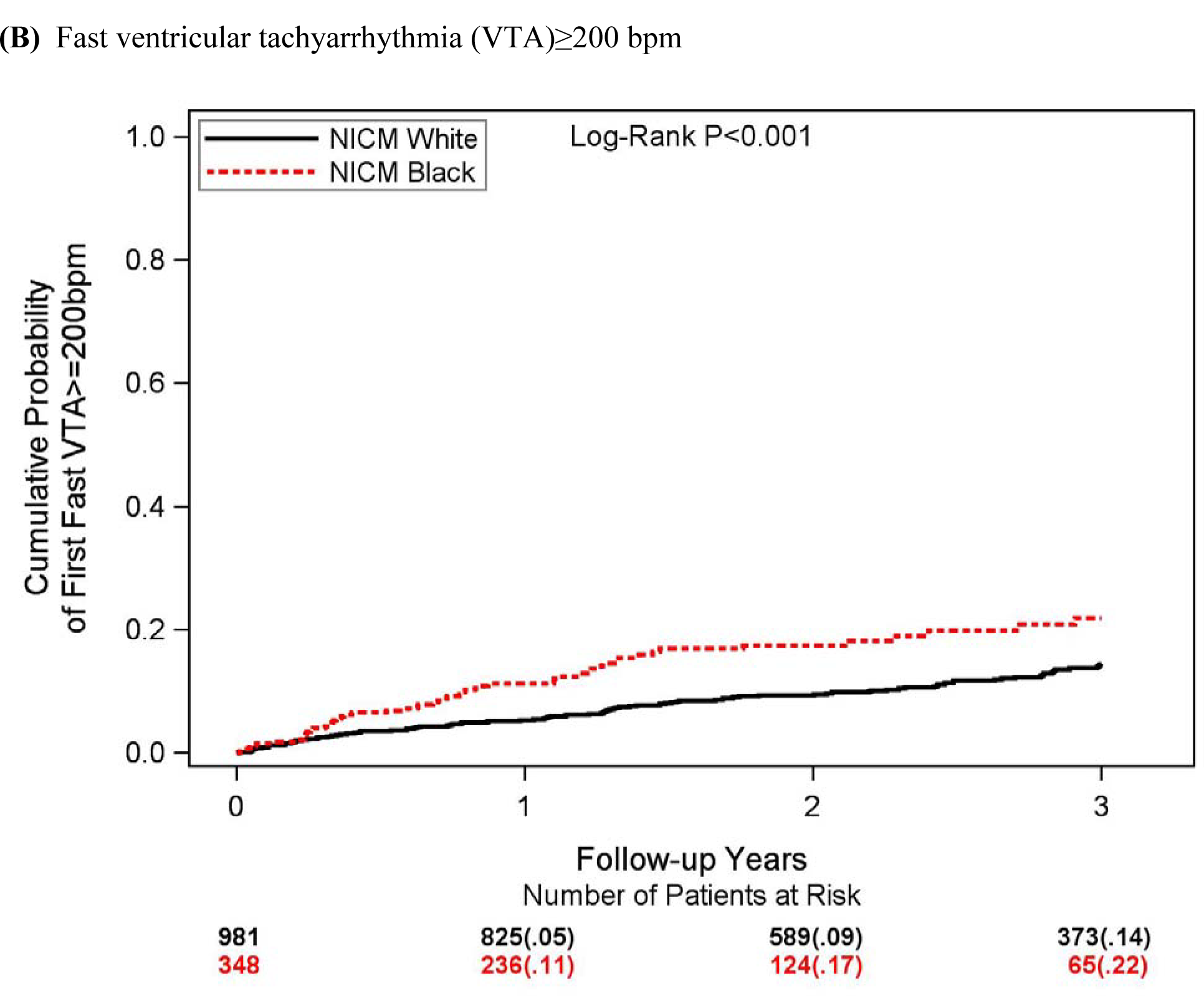
– Cumulative probability of ventricular tachyarrhythmia in NICM. 3-year Kaplan-Meier cumulative probability for (A) sustained ventricular tachyarrhythmia [VTA], and (B) fast VTA ≥200 bpm in Black and White patients with non-ischemic cardiomyopathy (NICM).

Similarly, the rate of first ATA, AF, and SVT were also significantly higher in Black compared with White patients (ATA 25% vs 12%; AF 9% vs 4%; SVT 21% vs 8%; respectively; p<0.001; Figure 2A-C).

**Figure 2.**
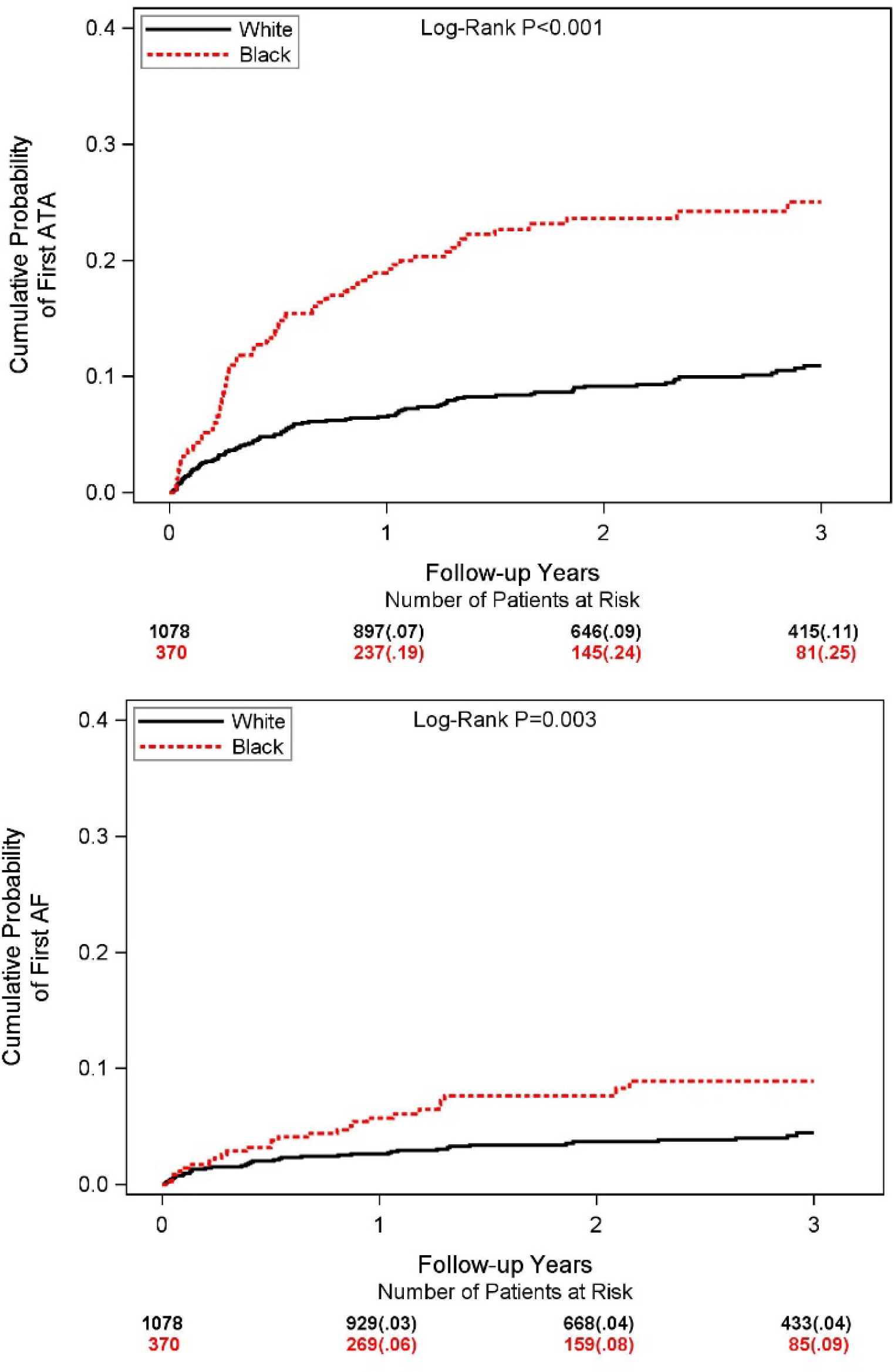
– Cumulative probability of ATA, AF, and SVT in NICM. 3-year Kaplan-Meier cumulative probability of; (A) atrial tachyarrhythmia (ATA), (B) atrial fibrillation (AF), and (C) supraventricular tachycardia (SVT) in Black and White patients with non-ischemic cardiomyopathy (NICM).

Consistent with the univariate findings, multivariable Cox modeling (Figure 3) adjusted for age, sex, smoking, diastolic blood pressure, CRT-D, NYHA class, HF hospitalization, and history of ATA, also showed that, compared with White patients with NICM, Black patients experienced a significantly higher risk of first VTA events/therapies, including: 77% higher risk of sustained VTA, 72% increased risk of fast VTA (≥200 bpm); and a 69% increased risk of a first appropriate ICD therapy (p<0.01 for all).

**Figure 3.**
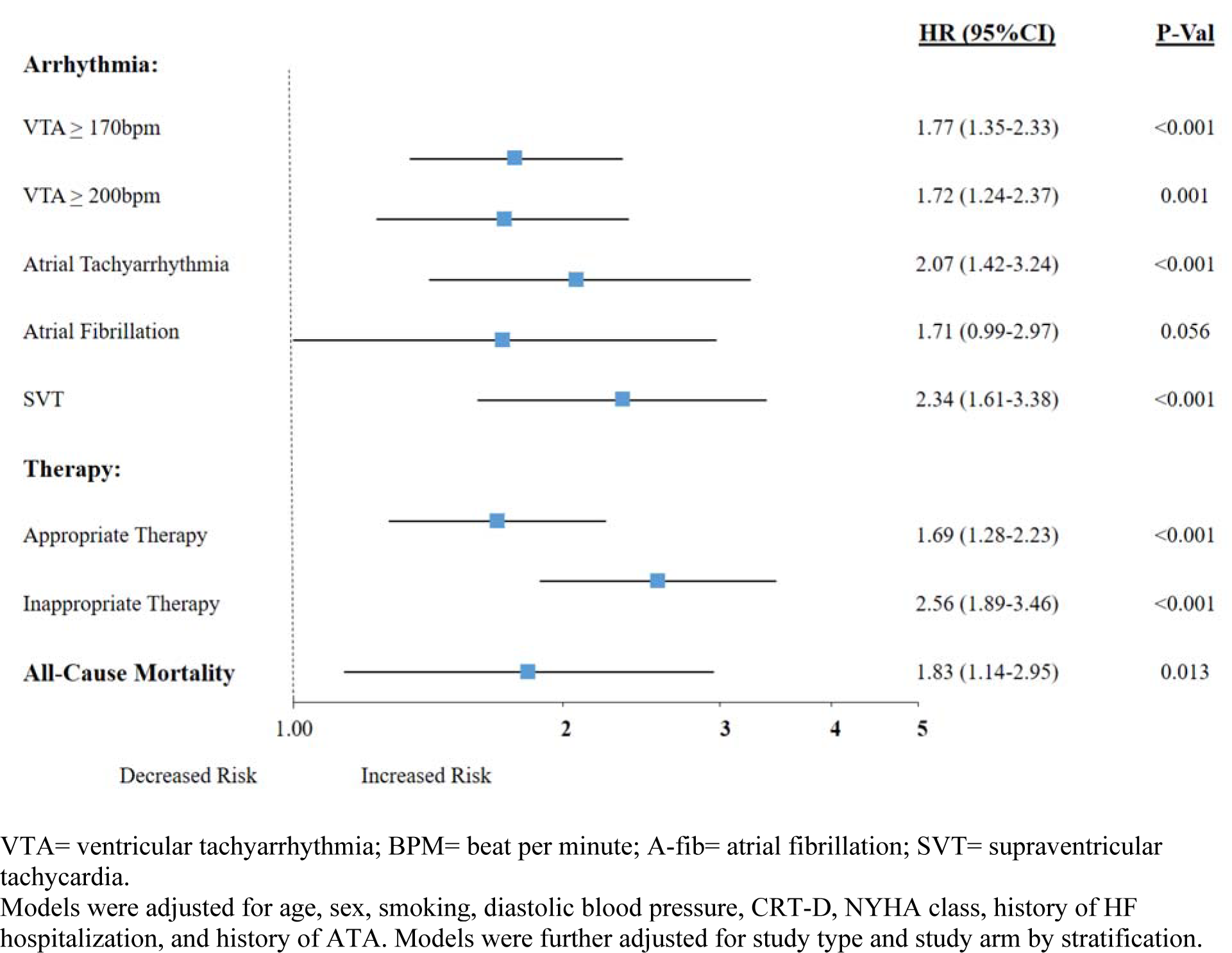
– Risk for first arrhythmic event, first ICD therapy, and death in NICM. Multivariable Cox regression evaluating the effect of Black vs. White on the development of arrhythmic endpoints, implantable cardioverter defibrillator (ICD) therapy, and death in patients with non-ischemic cardiomyopathy (NICM). VTA= ventricular tachyarrhythmia; BPM= beat per minute; A-fib= atrial fibrillation; SVT= supraventricular tachycardia. Models were adjusted for age, sex, smoking, diastolic blood pressure, CRT-D, NYHA class, history of HF hospitalization, and history of ATA. Models were further adjusted for study type and study arm by stratification.

Multivariate results were also consistent for ATA endpoints. Thus, compared with White patients, Blacks experienced a significant 107% increased risk for any ATA, 71% increased risk for AF, 134% increased risk for SVT, and a 156% increased risk for inappropriate therapy associated with ATA (Figure 3).

#### Recurrent events endpoints

Assessment of VTA burden showed similar findings. During follow-up, a total of 2581 VTA events occurred in 275 study patients with at least one event. At 3 years, the mean cumulative number of all types of arrhythmias/therapies was higher in Black than White patients (VTA: mean 1.0 vs. 0.55 events per patient [EPP]; Fast VTA: mean 0.43 vs 0.24 EPP; appropriate therapy: mean 0.43 vs 0.24 EPP; and inappropriate therapy: mean 0.43 vs 0.24 EPP; p<0.001; Figure 4A-B **and Supplementary Figure 3A-B**).

**Figure 4A-B.**
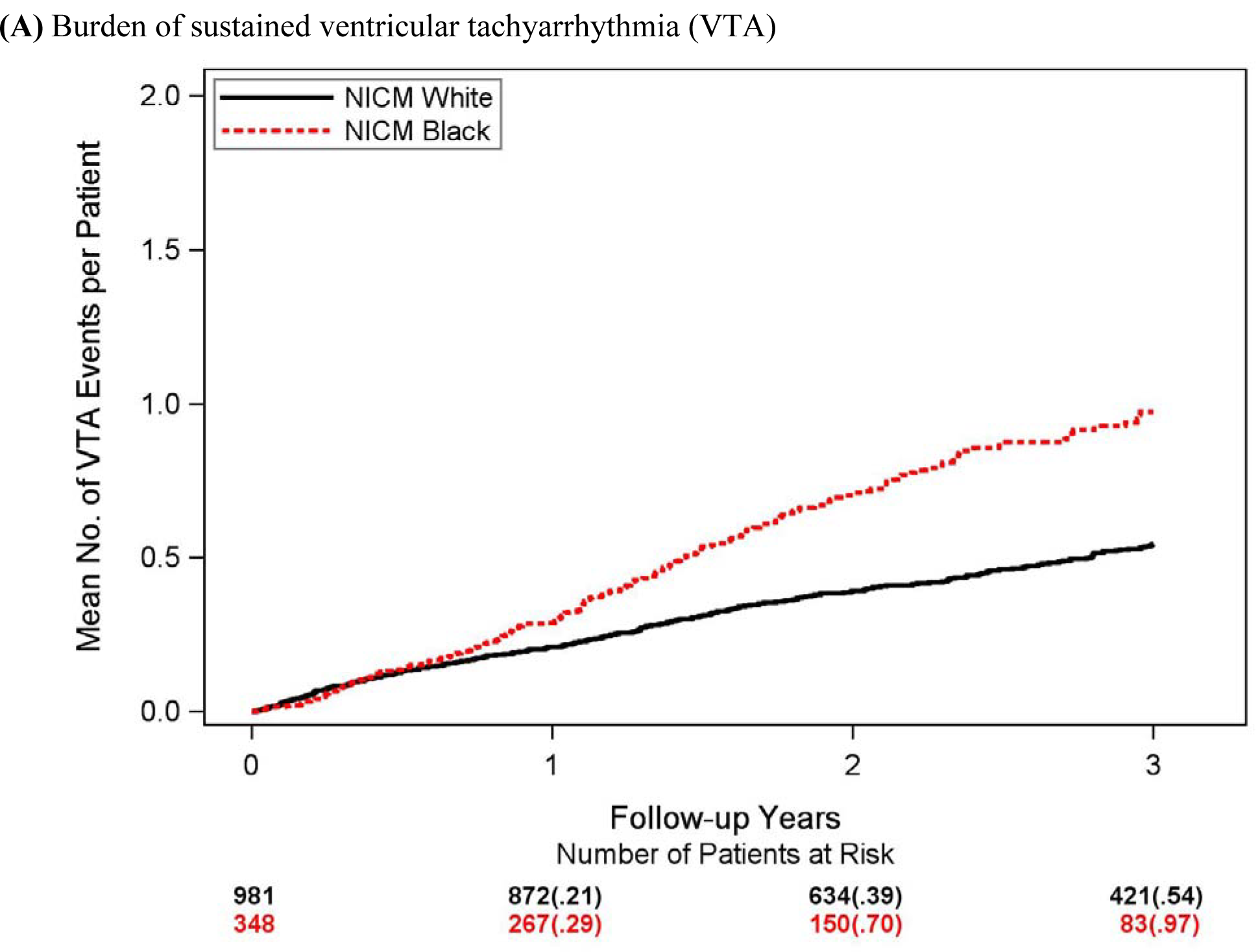
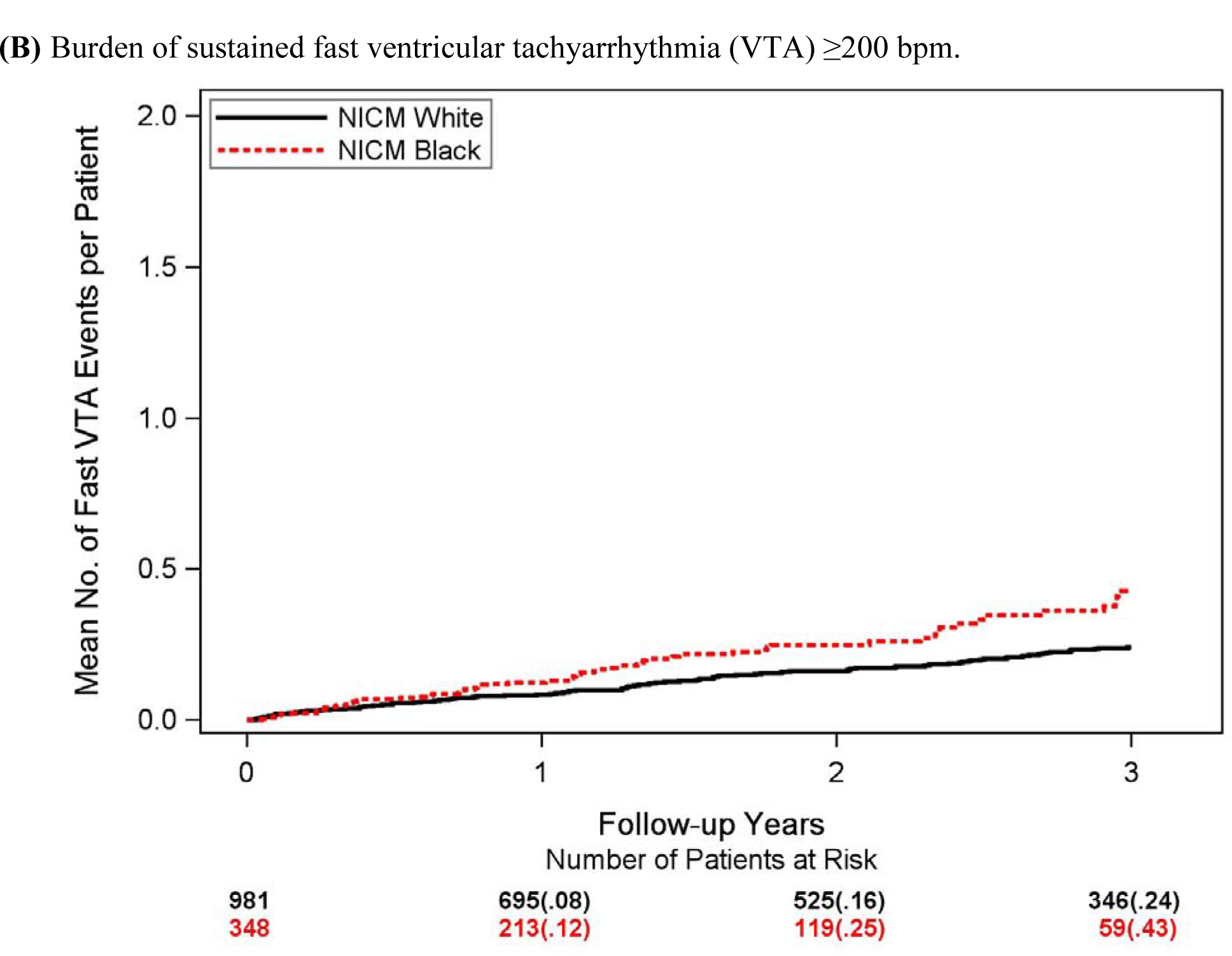
– The burden of ventricular tachyarrhythmia in <u>NICM</u>. Mean cumulative rate for recurrent events per patient stratified by study group for (A) ventricular tachyarrhythmia (VTA), and (B) fast VTA in Black and White patients with **<u>non-ischemic cardiomyopathy (NICM).</u>**

Multivariable analysis (**Supplementary Figure 4**) confirmed these findings and showed that, compared to White patients, Black patients exhibited a higher risk of high VTA burden (HR=1.84), fast VTA (HR=1.28), ATA (HR=2.59), appropriate therapies (HR=1.84), and inappropriate therapies (HR=2.90) (p<0.001 for all).

#### Subgroup analysis

Interaction-term analysis showed that the increased risk of VTA among Black vs. White patients with NICM was consistent regardless of age, sex, NYHA Class, the presence or absence of diabetes, the presence or absence of hypertension, and an ICD vs. CRT-D device (**Supplementary Appendix Table 3**).

#### Risk of death

Although Black patients were significantly younger (57±12 vs 62±12 years), they experienced a 2-fold higher rate of all-cause mortality at 3-years compared with White patients (12% vs 6%; p<0.001 for the overall comparison during follow-up; Figure 5). Cox proportional hazards regression analysis confirmed these findings after multivariate adjustment (HR=1.83; 95%CI 1.14-2.95; p=0.013; Figure 3).

**Figure 5.**
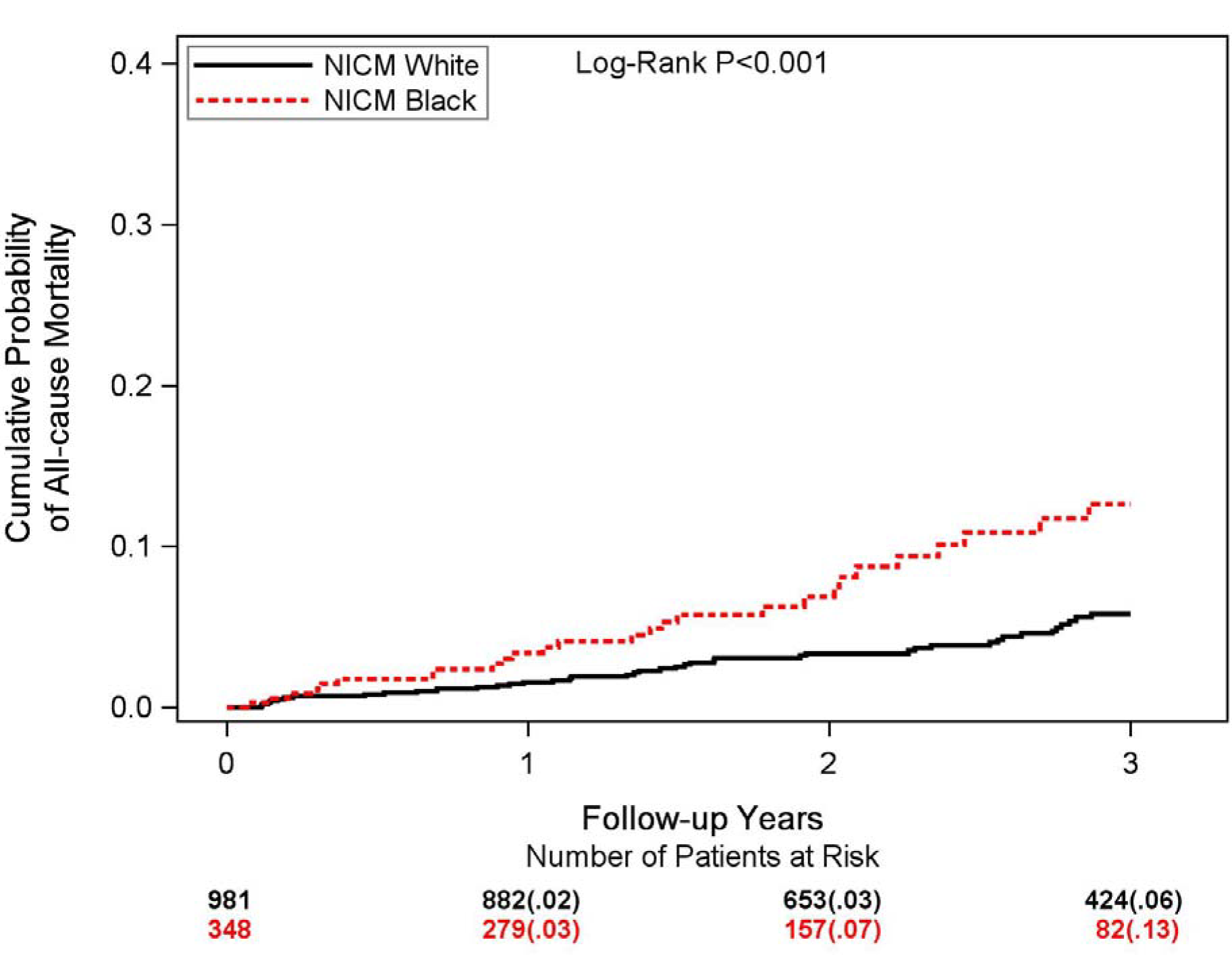
– Cumulative probability of all-cause mortality in NICM. 3-year Kaplan-Meier cumulative probability of all-cause mortality in Black and White patients with non-ischemic cardiomyopathy.

### Patients with ischemic cardiomyopathy

At 3 years, the Kaplan Meier cumulative probability of a first VTA was similar among Black and White patients (22% vs. 23%, respectively; p=0.55 for the overall difference during follow-up; **Supplementary Figure 5**). Similarly, the cumulative rate of first episode of fast VTA (19% vs. 14%; p=0.15), appropriate ICD therapy (25% vs. 23%; p=0.53), and inappropriate therapy (12% vs. 11%; p=0.28) did not differ significantly between Black and White patients (**Supplementary Figure 5B-D**).

Assessment of VTA burden showed similar findings. At 3 years, the mean cumulative number of all types of arrhythmias/therapies did not differ significantly between Black and White patients (**Supplementary Figure 6A-D**).

The cumulative rate of all-cause mortality was also similar between Black and White patients despite Black being implanted at a younger age (15% vs. 16%; p=0.42; **Supplementary Figure 7**).

## Discussion

Our study utilizing a combined database of ICM and NICM patients enrolled in MADIT Trials and the recent RAID trial addresses disparities in the risk of arrhythmic events and survival between Black and White patients with an ICD for primary prevention. Our findings show that in patients with NICM, self-identified Black patients compared with White patients have many differences including 1) a significantly younger age of presentation and higher burden of comorbidities; 2) a significantly higher incidence and burden of VTA, and ATA events; 3) increased incidence and burden of appropriate and inappropriate ICD therapies; and 4) lower survival rates, despite receiving an ICD at a significantly younger age. In contrast, in patients with ICM, the risk of tachyarrhythmias and ICD therapies did not differ significantly between Black and White patients. Our findings highlight the important disparities between Black and White patients with a NICM, suggesting a need for more intensive arrhythmia monitoring and management, as well as potentially earlier consideration for more advanced HF therapies in Black patients due to increased likelihood for device therapy, adverse arrhythmic events, and mortality following prophylactic ICD implantation.

### Disparities in outcomes between Black and White patients with NICM

In the U.S., Black patients have a higher prevalence of NICM at a younger age than the general population. The exact reason for this disparity is not fully understood, but potential contributing factors may include differences in the prevalence of comorbidities associated with NICM, environmental exposures, healthcare access, as well as potential genetic factors.^11, 12, 20^ Furthermore, Black patients with NICM frequently exhibit unique clinical presentations and imaging findings that differ from other populations. For example, Black patients with NICM are more likely to present with HF symptoms at an earlier age and with a higher burden of myocardial fibrosis on cardiac MRI.^21, 22^

Data on clinical outcomes suggest that Black patients with NICM have a higher risk of HF hospitalization and mortality compared to White patients with NICM.^23–26^ Differences in arrhythmic burden remained controversial due to under-representation of Blacks in randomized HF ICD trials.^27–30^ Our meta-analysis utilizing the largest cohort of Black patients showed a significantly higher risk of tachyarrhythmias (VTA and or ATA), ICD therapies (appropriate and inappropriate), as well as all-cause and non-arrhythmic mortality among Black patients with NICM when compared with White patients even after adjusting for age, sex, NYHA class, diabetes, and hypertensions. These findings suggest a need for close arrhythmia monitoring and consideration of early intervention in Black patients with NICM regardless of baseline risk factors.

### Arrhythmic outcomes among Black and White patients with ICM

Prior data on outcomes of Black patients in the ICM group were inconsistent. The majority of the findings from real world data are derived from studies that did not assess outcomes by the type of cardiomyopathy, and show increased risk of SCD, all-cause mortality, and tachyarrhythmia (VTA and/or ATA) among the Black patients when compared with White patients.^31–34^ On the other hand, subgroup analysis in ICM patients only (N=642) enrolled in the Prospective Observational Study of Implantable Cardioverter-Defibrillators (PROSE-ICD) showed similar risks for ICD therapy, and all-cause mortality between Black and White.^28^ The landmark ICD trials MUSST, DEFINITE and DINAMIT included only a minority of Black patients and were underpowered to conduct valid analysis. The SCD-HeFT trial showed similar findings between Black and White, however subgroup analysis looking at ICM patients only was not performed.^27^ Our data are in line with these observations, and show that in patients with ICM who are implanted with an ICD in the U.S., arrhythmic and mortality outcomes are similar Black and White patients.

### Possible mechanisms underlying observed difference in arrhythmic risk between Black and White patients by type of cardiomyopathy

Several studies have suggested that the increased arrhythmic risk seen in Black patients may be attributed to social determinants of health. However, the fact that these differences are dependent on type of heart disease makes this less likely. Results from the Atherosclerosis Risk in Communities Study (ARIC) which included 3,832 Black patients and 11,237 Whites patients showed that income, education, and traditional risk factors explained less than 65% of the difference in SCD.^33^ Although the mechanism for differences in arrhythmic burden between Black and White patients remains unknown, it is likely multifactorial. Potential mechanisms include differences in baseline comorbidities, social determinants of health, environmental, and possibly genetic factors that affect response to medical therapies, such as angiotensin-converting enzyme inhibitors and or beta-blockers.^35–37^ In contrast to ICM, where VTA is mostly scar related (which supposed to be similar in Black and White patients), NICM consists of a heterogeneous group of cardiomyopathies, and the distribution of the specific cardiomyopathies within this group is known to be different between Black and White patients.^38, 39^

### Management implications

Regardless of potential mechanisms underlying these disparities in arrhythmic outcomes among patients with NICM, our findings show that Black patients experienced both increased VTA and mortality risk compared with White patients. These findings have important implications with regard to the potential benefit of the ICD in this population. The predicted benefit of the ICD can be calculated as the difference in the risk for fast VTA and the risk for death without a prior fast VTA in life.^40^ As seen in **Figure 8A-B in the Supplementary Appendix**, this difference was similar and pronounced in both Black and White patients. These findings highlight the importance of early intervention with a prophylactic ICD in Black patients with NICM due to increased risk of VTA in this population, as well as careful follow-up and consideration of early intensification of therapies following ICD implantation, due to the increased burden of appropriate and inappropriate therapies and lower survival rates in Black patients with NICM who receive an ICD.

In the U.S., HF patients who identify themselves as Black are less likely to be referred to specialists, less likely to receive ICD counseling, and less likely to have an ICD implanted when compared to White HF patients.^41–43^ These factors contribute to under-utilization of ICDs in Black patients and ultimately to disparities in survival outcomes.

### Limitations

Our study is subject to several limitations. The current analysis is a post-hoc and non-pre-specified analysis; thus, it should be considered a hypothesis-generating. Although our models were adjusted for significant predictors, which helps reduce uncertainty when estimating the association between Black and the hazard of VTA/death, it is still possible that the observed differences in arrhythmic and survival outcomes between Black and White patients in the present study were driven by differences in the frequency of comorbidities and other social determinants of health between Black and White patients that were not accounted for the multivariable models.

In addition, the eligibility criteria and the exclusion of elderly patients or those with advanced diseases (such as patients with creatinine >2.5 mg/dL) in all our trials limits the generalizability of these findings to these populations. The impact of very recent HF drugs (including angiotensin receptor neprilysin inhibitor and Sodium glucose co-transporter 2 inhibitors) on the VTA risk in ICD candidates remains unknown. Add also a note that the findings of this study pertain only to Black patients who are implanted with an ICD in the U.S. that may be affected by disparities and SDOH that may not be present in other countries.,

## Conclusions and clinical implications

Our combined data from landmark ICD trials, employing uniform arrhythmia and mortality adjudication data, show that in NICM Black patients experience increased risk and burden of tachyarrhythmias, ICD therapies, as well as lower survival rates following ICD implant when compared to White patients. These disparities in arrhythmic and clinical outcomes suggest the need for early intervention with an ICD, careful monitoring, and intensification of HF and antiarrhythmic therapies, in Black patients with NICM in the U.S.

## Data Availability

Data is available upon request only.

## Acknowledgment

The following authors Arwa Younis, Shireen Saxena, Ilan Goldenberg, Scott McNitt, and Bronislava Polonsky had full access to all the data in the study and take responsibility for the integrity of the data and the accuracy of the data analysis.

Each of the MADIT trials were funded by an unrestricted research grant from Boston Scientific to the University of Rochester Medical Center, Rochester, NY. Nevertheless, the design and conduct of the study; collection, management, analysis, and interpretation of the data; preparation, review, or approval of the manuscript; and decision to submit the manuscript for publication was made by the authors only.

---

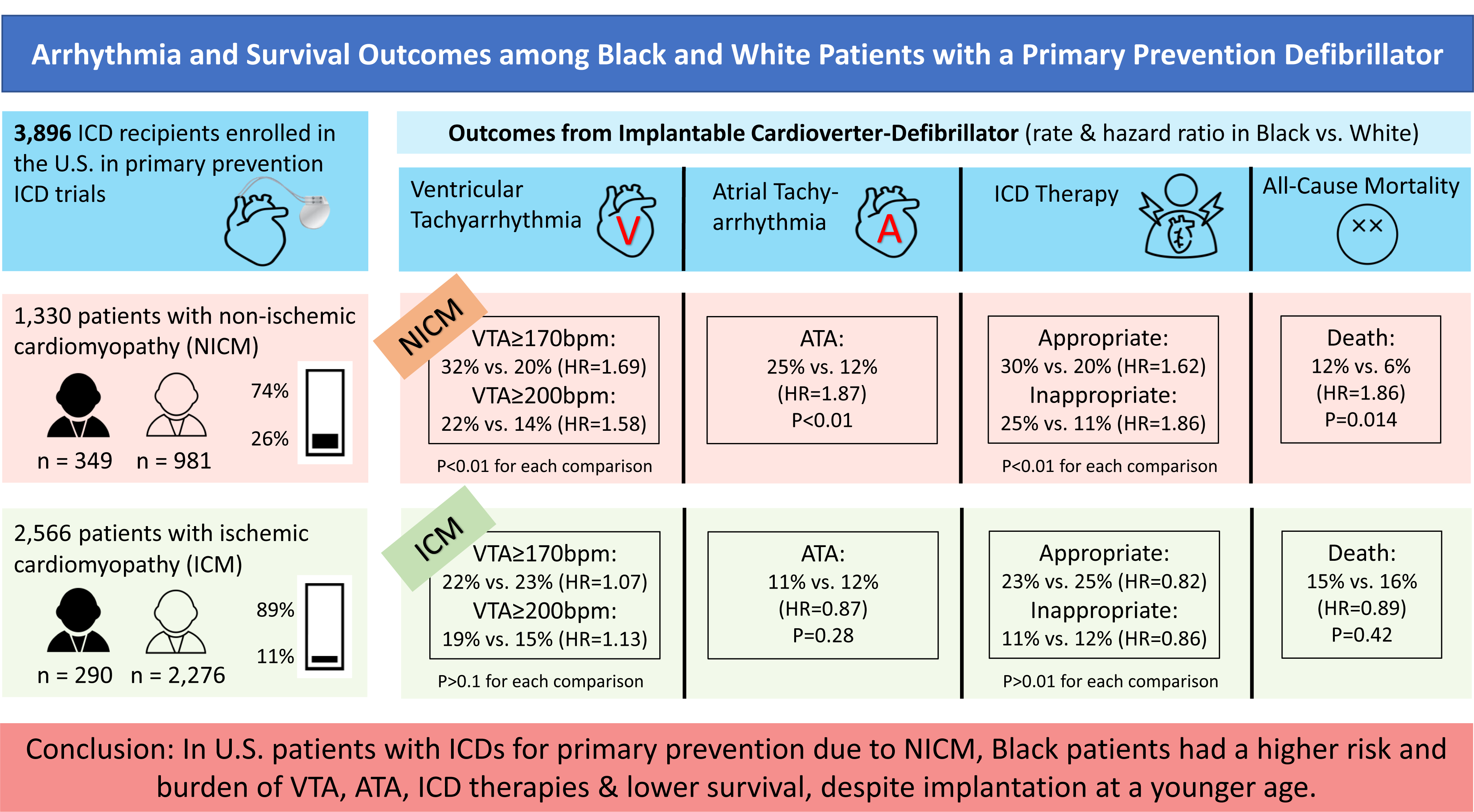

## References

1. Sheikh FH, Ravichandran AK, Goldstein DJ, Agarwal R, Ransom J, Bansal A, Kim G, Cleveland JC, Uriel N, Sheridan BC, Chomsky D, Patel SR, Dirckx N, Franke A and Mehra MR. Impact of Race on Clinical Outcomes After Implantation With a Fully Magnetically Levitated Left Ventricular Assist Device: An Analysis From the MOMENTUM 3 Trial. Circulation Heart failure. 2021;14:e008360.

2. Truby LK and Rogers JG. Advanced Heart Failure: Epidemiology, Diagnosis, and Therapeutic Approaches. JACC: Heart Failure. 2020;8:523-536.

3. Cuyjet AB and Akinboboye O. Acute Heart Failure in the African American Patient. Journal of Cardiac Failure. 2014;20:533-540.

4. Colantonio LD, Gamboa CM, Richman JS, Levitan EB, Soliman EZ, Howard G and Safford MM. Black-White Differences in Incident Fatal, Nonfatal, and Total Coronary Heart Disease. 2017;136:152-166.

5. Smilowitz NR, Maduro GA, Jr., Lobach IV, Chen Y and Reynolds HR. Adverse Trends in Ischemic Heart Disease Mortality among Young New Yorkers, Particularly Young Black Women. PloS one. 2016;11:e0149015.

6. Safford MM, Brown TM, Muntner PM, Durant RW, Glasser S, Halanych JH, Shikany JM, Prineas RJ, Samdarshi T, Bittner VA, Lewis CE, Gamboa C, Cushman M, Howard V and Howard G. Association of race and sex with risk of incident acute coronary heart disease events. Jama. 2012;308:1768-74.

7. Feng Qian CSP, Sarwart I. Chaudhry, Edward L. Hannan, Benjamin A. Shaw, John A. Spertus, Harlan M. Krumholz. Racial Differences in Heart Failure Outcomes JACC: Heart Failure. 2015;3:531-538.

8. Yevgeniy Khariton MEN, Laine Thomas, Gregg C. Fonarow, Xiaojuan Mi, Adam D. Devore, Carol Duffy, Puza P. Sharma, Nancy M. Albert, J. Herbert Patterson, Javed Butler, Adrian F. Hernandez, Fredonia B. Williams, Kevin McCague, John A. Spertus Health Status Disparities by Sex, Race/Ethnicity, and Socioeconomic Status in Outpatients with Heart Failure. JACC: Heart Failure. 2018;6:465-473.

9. Petery Glynn DML-J, Matthew J. Feinsetin, Merecedes Carnethon, Sadiya S. Khan. Disparities in Cardiovascular Mortality Related to Heart Failure in the United States. Journal of the American College of Cardiology 2019;73:2354-2355.

10. Samul T. Savitz TL, Sue Hee Sung, Keane Lee, Jamal S. Rana, Grace Tabada, Alan S. Go. Contemporary Reevaluation of Race and Ethnicity with Outcomes in Heart Failure. Journal of the American Heart Association 2021;10.

11. Coughlin SS, Myers L and Michaels RK. What explains black-white differences in survival in idiopathic dilated cardiomyopathy? The Washington, DC, Dilated Cardiomyopathy Study. Journal of the National Medical Association. 1997;89:277-82.

12. Coughlin SS, Labenberg JR and Tefft MC. Black-white differences in idiopathic dilated cardiomyopathy: the Washington DC dilated Cardiomyopathy Study. Epidemiology (Cambridge, Mass). 1993;4:165-72.

13. Musemwa N and Gadegbeku CA. Hypertension in African Americans. Current cardiology reports. 2017;19:129.

14. Gaskin DJ, Thorpe RJ, Jr., McGinty EE, Bower K, Rohde C, Young JH, LaVeist TA and Dubay L. Disparities in diabetes: the nexus of race, poverty, and place. American journal of public health. 2014;104:2147-55.

15. Haw JS, Shah M, Turbow S, Egeolu M and Umpierrez G. Diabetes Complications in Racial and Ethnic Minority Populations in the USA. Current diabetes reports. 2021;21:2.

16. Moss AJ, Zareba W, Hall WJ, Klein H, Wilber DJ, Cannom DS, Daubert JP, Higgins SL, Brown MW and Andrews ML. Prophylactic implantation of a defibrillator in patients with myocardial infarction and reduced ejection fraction. The New England journal of medicine. 2002;346:877-83.

17. Moss AJ, Hall WJ, Cannom DS, Klein H, Brown MW, Daubert JP, Estes NA, 3rd, Foster E, Greenberg H, Higgins SL, Pfeffer MA, Solomon SD, Wilber D, Zareba W and Investigators M-CT. Cardiac-resynchronization therapy for the prevention of heart-failure events. N Engl J Med. 2009;361:1329-38.

18. Moss AJ, Schuger C, Beck CA, Brown MW, Cannom DS, Daubert JP, Estes NA, 3rd, Greenberg H, Hall WJ, Huang DT, Kautzner J, Klein H, McNitt S, Olshansky B, Shoda M, Wilber D and Zareba W. Reduction in inappropriate therapy and mortality through ICD programming. The New England journal of medicine. 2012;367:2275-83.

19. Zareba W, Daubert JP, Beck CA, Huang DT, Alexis JD, Brown MW, Pyykkonen K, McNitt S, Oakes D, Feng C, Aktas MK, Ayala-Parades F, Baranchuk A, Dubuc M, Haigney M, Mazur A, McPherson CA, Mitchell LB, Natale A, Piccini JP, Raitt M, Rashtian MY, Schuger C, Winters S, Worley SJ, Ziv O and Moss AJ. Ranolazine in High-Risk Patients With Implanted Cardioverter-Defibrillators: The RAID Trial. Journal of the American College of Cardiology. 2018;72:636-645.

20. Coughlin SS, Neaton JD, Sengupta A and Kuller LH. Predictors of mortality from idiopathic dilated cardiomyopathy in 356,222 men screened for the Multiple Risk Factor Intervention Trial. American journal of epidemiology. 1994;139:166-72.

21. Arabadjian ME, Yu G, Sherrid MV and Dickson VV. Disease Expression and Outcomes in Black and White Adults With Hypertrophic Cardiomyopathy. 2021;10:e019978.

22. Francone M. Role of cardiac magnetic resonance in the evaluation of dilated cardiomyopathy: diagnostic contribution and prognostic significance. ISRN radiology. 2014;2014:365404.

23. Bozkurt B, Colvin M, Cook J, Cooper LT, Deswal A, Fonarow GC, Francis GS, Lenihan D, Lewis EF, McNamara DM, Pahl E, Vasan RS, Ramasubbu K, Rasmusson K, Towbin JA and Yancy C. Current Diagnostic and Treatment Strategies for Specific Dilated Cardiomyopathies: A Scientific Statement From the American Heart Association. 2016;134:e579-e646.

24. Brown DW, Haldeman GA, Croft JB, Giles WH and Mensah GA. Racial or ethnic differences in hospitalization for heart failure among elderly adults: Medicare, 1990 to 2000. American heart journal. 2005;150:448-54.

25. Can AD and Coughlin SS. Addressing Health Disparities in Idiopathic Dilated Cardiomyopathy. Jacobs journal of community medicine. 2017;3.

26. Coughlin SS. Toward the primary prevention of idiopathic dilated cardiomyopathy. Journal of the National Medical Association. 1991;83:949-50.

27. Mitchell JE, Hellkamp AS, Mark DB, Anderson J, Poole JE, Lee KL and Bardy GH. Outcome in African Americans and other minorities in the Sudden Cardiac Death in Heart Failure Trial (SCD-HeFT). American heart journal. 2008;155:501-6.

28. Zhang Y, Kennedy R, Blasco-Colmenares E, Butcher B, Norgard S, Eldadah Z, Dickfeld T, Ellenbogen KA, Marine JE, Guallar E, Tomaselli GF and Cheng A. Outcomes in African Americans undergoing cardioverter-defibrillator implantation for primary prevention of sudden cardiac death: findings from the Prospective Observational Study of Implantable Cardioverter-Defibrillators (PROSE-ICD). Heart Rhythm. 2014;11:1377-83.

29. Kyndaron Reiner GAN, Adriana Huertas-Vasquez, Audrey Uy-Evanado, Carmen Teodorescu, Eric C Stecker, Karen Gunson, Jonathan Jui, Sumeet S Chugh Distinctive Clinical Profiles of Blacks versus Whites Presenting with Sudden Cardiac Arrest. Circulation 2015;132:380-387.

30. Avi Sabbag IG, Arthur Moss, Scott McNitt, Slava Polonsky, Wojciech Zareba, Valentina Kutyifa Abstract 17223: The risk of Ventricular Tachyarrthymias or Death in Black or White Cardiac Patients - Madit-CRT sub study. Circulation. 2018;130.

31. Groeneveld PW, Heidenreich PA and Garber AM. Racial disparity in cardiac procedures and mortality among long-term survivors of cardiac arrest. Circulation. 2003;108:286-91.

32. Thomas KL, Al-Khatib SM, Kelsey RC, 2nd, Bush H, Brosius L, Velazquez EJ, Peterson ED and Gilliam FR. Racial disparity in the utilization of implantable-cardioverter defibrillators among patients with prior myocardial infarction and an ejection fraction of &lt;or=35%. The American journal of cardiology. 2007;100:924-9.

33. Zhao D, Post WS, Blasco-Colmenares E, Cheng A, Zhang Y, Deo R, Pastor-Barriuso R, Michos ED, Sotoodehnia N and Guallar E. Racial Differences in Sudden Cardiac Death. 2019;139:1688-1697.

34. Jain V, Minhas AMK, Kleiman NS, Arshad HB, Saleh Y, Pandat SS, Dani SS, Goel SS, Faza N, Butt SA, Blankstein R, Cainzos-Achirica M, Nasir K and Khan SU. Cardiac Arrest in Young Adults With Ischemic Heart Disease in the United States, 2004-2018. Current problems in cardiology. 2022;47:101312.

35. Lanfear DE, Hrobowski TN, Peterson EL, Wells KE, Swadia TV, Spertus JA and Williams LK. Association of β-blocker exposure with outcomes in heart failure differs between African American and white patients. Circulation Heart failure. 2012;5:202-8.

36. Cresci S, Kelly RJ, Cappola TP, Diwan A, Dries D, Kardia SL and Dorn GW, 2nd. Clinical and genetic modifiers of long-term survival in heart failure. Journal of the American College of Cardiology. 2009;54:432-44.

37. Breathett K, Leng I, Foraker RE, Abraham WT, Coker L, Whitfield KE, Shumaker S, Manson JE, Eaton CB, Howard BV, Ijioma N, Cené CW, Martin LW, Johnson KC, Klein L, Rossouw J, Ludlam S, Burwen D, McGowan J, Ford L, Geller N, Anderson G, Prentice R, LaCroix A, Kooperberg C, Stefanick ML, Jackson R, Thomson CA, Wactawski-Wende J, Limacher M, Wallace R and Kuller L. Risk Factor Burden, Heart Failure, and Survival in Women of Different Ethnic Groups. 2018;11:e004642.

38. Mirsaeidi M, Machado RF, Schraufnagel D, Sweiss NJ and Baughman RP. Racial difference in sarcoidosis mortality in the United States. Chest. 2015;147:438-449.

39. Gentry MB, Dias JK, Luis A, Patel R, Thornton J and Reed GL. African-American women have a higher risk for developing peripartum cardiomyopathy. Journal of the American College of Cardiology. 2010;55:654-9.

40. Younis A, Goldberger JJ, Kutyifa V, Zareba W, Polonsky B, Klein H, Aktas MK, Huang D, Daubert J, Estes M, Cannom D, McNitt S, Stein K and Goldenberg I. Predicted benefit of an implantable cardioverter-defibrillator: the MADIT-ICD benefit score. European heart journal. 2021;42:1676-1684.

41. Hess PL, Hernandez AF, Bhatt DL, Hellkamp AS, Yancy CW, Schwamm LH, Peterson ED, Schulte PJ, Fonarow GC and Al-Khatib SM. Sex and Race/Ethnicity Differences in Implantable Cardioverter-Defibrillator Counseling and Use Among Patients Hospitalized With Heart Failure: Findings from the Get With The Guidelines-Heart Failure Program. Circulation. 2016;134:517-26.

42. El-Chami MF, Hanna IR, Bush H and Langberg JJ. Impact of race and gender on cardiac device implantations. Heart Rhythm. 2007;4:1420-6.

43. Hernandez AF, Fonarow GC, Liang L, Al-Khatib SM, Curtis LH, LaBresh KA, Yancy CW, Albert NM and Peterson ED. Sex and racial differences in the use of implantable cardioverter-defibrillators among patients hospitalized with heart failure. Jama. 2007;298:1525–32.

